# Prevalence and Outcome of Adults with Congenital Heart Disease and Coronary Artery Disease

**DOI:** 10.1101/2023.06.05.23290998

**Authors:** Haiyang Li, Yan Yan, Shipan Wang, Mingyu Sun, Sichong Qian, Yuan Xue, Hongjia Zhang

## Abstract

**BACKGROUND:** Little is known about the prevalence of outcome in patients with coronary artery disease (CAD) among adults with congenital heart disease (ACHD). The objectives of this study were to determine rates of ACHD and factors associated with outcomes among adults with CAD in a contemporary cohort within the China.

**METHODS:** Using pragmatic data from the Prospective Registry of the Current Status of Care for Patients with Coronary Artery Disease database, between January and May 2022, we stratified all CAD patients with comorbid ACHD or not. Using logistic regression method, we explored the risk factors associated with lower risk of mortality and stroke. We constructed a matching sub-cohort as sensitive analysis.

**RESULTS:** Of 10,517 evaluable patients 189 (1.8%) were with a diagnosis of ACHD, and were more female, more frequent with a history of atrial fibrillation, had a lower hemoglobin level at admission but less likely to have hypertension or smoking history than those without ACHD. In-hospital mortality before discharge was higher in patients with ACHD compared with no ACHD in the whole population (1.06% vs. 0.28%, p=0.05) and matched population (1.06% vs. 0.13%, p=0.04). Acute myocardial infarction (54.8%) represented the most common cause of death among all cohort, whereas 100% deaths among ACHD were due to ischemic stroke (2 cases).

**CONCLUSIONS**

ACHD was associated with a higher rate of mortality, which stroke complications contribute most. Future studies are needed to improve the acknowledge of underlying stroke mechanisms and prognosis in this special population.

## INTRODUCTION

Given the success of modern cardiology care, congenital heart disease (CHD) has been transformed from a deadly condition into a treatable disorder, with an increasing need to deliver age-appropriate medical care^1^. Numerous studies have demonstrated that residual defects contribute substantially to long-term morbidity^2–4^. The current Guidelines for Adults with Congenital Heart Disease (ACHD) developed for general cardiologists and the community of ACHD specialists focusing on surgical and percutaneous interventions process^5, 6^. However, somewhat surprising is the fact that recommendations have largely been classified as Class I with a level of evidence at B or C, indicating the mismatch between clinical needs and quality of supportive data^5^.

As a field which lags significantly behind other adult populations, specialized study performing on ACHD are warranted to generate comparable evidence to non-CHD population. In the present study, we aimed to provide a general overview of ACHD in the coronary artery disease (CAD) population using a pragmatic registry. We also aim to provide an outlook on the potential prognostic factors and outcomes in this population.

## METHODS

### Study design and patient selection

Data from the Prospective Registry of the Current Status of Care for Patients with Coronary Artery Disease database were used in the study. Launched on January 17th, 2022, the project (clinicaltrial.gov NCT 05337319) is a nationwide, pragmatic registry including patients with a principal discharge diagnosis of coronary artery disease (CAD). The registry is a program of the National Clinical Research Center for Cardiovascular Disease in Beijing Anzhen hospital, focusing on quality improvement efforts for CAD care by routinely collected electronic health records. Briefly, the in-hospital patients were recruited consecutively in collaborative hospitals every month. Patient were eligible for enrollment with a principal discharge diagnosis of CAD, including ST elevated myocardial infarction, non-ST-segment elevation myocardial infarction, unstable angina pectoris, and stable coronary artery disease. The protocol was approved by the Ethics Committee of Beijing Anzhen Hospital, Capital Medical University.

Patient-level data including demographics, medical history, physical examination, laboratory findings, medical treatment, procedural details, and in-hospital outcomes were collected via electronic medical records (EMR) and death registry system databases. Diagnosis and procedural details were entered into the database by medical staffs and coded by trained data abstractors. Data quality including accuracy, completeness, and consistency was monitored monthly by the study management group. All consecutive in-hospital patients with CAD at discharge in the database between January 17th, 2022 and May 31th, 11023 were included in this study. Patients who were not older than 18 years were excluded from the analysis.

### Definitions of variables

ICD-10 diagnosis codes and ICD-9-CM-3 procedural codes were utilized for variable collection^7, 8^. CAD and CHD were defined in accordance with the guidelines published by the Chinese society of cardiology and recorded by doctors on original medical records^9^. Patients were considered to have CHD, such as atrial septal defect, ventricular septal defect, or patent ductus arteriosus, if those had been documented in admission or detected by echocardiogram during hospitalization^10^. An estimated glomerular filtration rate lower than 60 mL/min per 1.73 m^2^ was defined as renal insufficiency. Patients were classified as ACHD group if they had firmed evidence of ACHD during their hospitalization for treating CAD. Patients who were not recognized any congenital cardiac defect were defined as “no ACHD”.

### Endpoints

The primary endpoint was in-hospital mortality. Cardiovascular death was defined as any death with a clear relationship to underlying coronary heart disease (including death, secondary to acute MI, sudden death, unobserved and unexpected death, resuscitated out of hospital cardiac arrest that does not survive to hospital discharge), stroke, and other death that cannot definitely be attributed to a non-CV cause. Stroke was defined as a new neurologic deficit during the index event hospitalization. Both ischemic and hemorrhagic stroke will be considered as an endpoint and differentiated into the 2 types of events. Pneumonia was defined as new documentation in the medical record based on supportive laboratory findings and radiologic evidence. All these endpoints were documented in medical records by clinical doctors during hospitalization.

### Statistical analysis

Baseline characteristics and procedural details of included patients are described. Continuous variables are presented as mean and standard deviation (SD) if a normal distribution, and as a median and interquartile range (IQR) if skewed. Categorical variables are presented as frequency and percentage. T-test or Kruskal-Wallis test for continuous variables, and Chi-square test for categorical variables were used to test for statistically significant differences between the ACHD versus No ACHD groups.

We used logistic regression method to explore the potential risk factors of endpoints. The univariate logistic models were built by each candidate that showed a significant difference between the ACHD and no ACHD groups. Odd ratios (OR) and 95% confidence intervals (CI) were calculated and reported.

Considering the unbalanced number of enrolled patients between two groups, we constructed a matching sub-cohort from this population to conduct the sensitive analysis. In detail, we used all ACHD patients as ‘cases’ and selected 4 ‘controls’ for each of them from the No ACHD patients randomly. Each control had a same age and sex with the matched case. We applied logistic models in this matched cohort to verify the association between potential risk factors and endpoints.

The reference was used as No ACHD group in all analyses. All tests were two-sided, and a *p* value of less than 0.05 was considered with statistical significance. All statistical analyses were performed by SAS version 9.4 (SAS Institute Inc; Cary, NC).

## RESULTS

### Patient characteristics, treatments, and procedural details

Up to May 31, 2022, a total of 11,023 patients with cleaned data were enrolled in the study. Based on the principal discharge diagnosis, 10,519 patients with CAD during the study period were identified. After excluding 2 patients younger than 18 years, 10,517 patients were included in the study, of whom 189 (1.8%) were diagnosed with ACHD. The type of CHD in ACHD group is shown in **Figure 1**. 750 No ACHD patients were selected as controls (6 ACHD patients could only find 3 controls for each).

**Figure 1.**
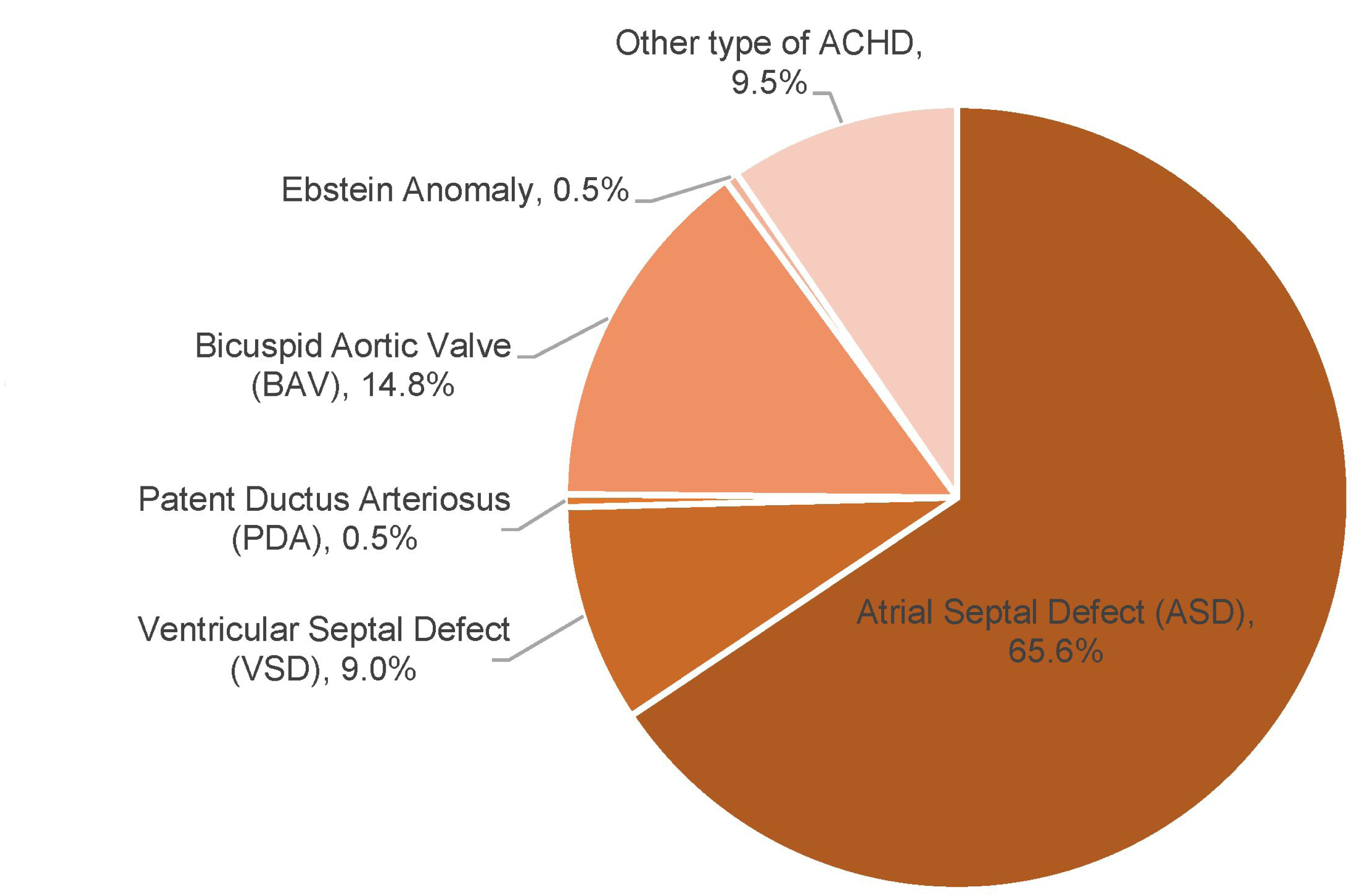
Type of CHD in ACHD group. CHD, congenital heart disease; ACHD, congenital heart disease

Patient baseline characteristics and procedural details were shown in **Table 1**. Patients with ACHD and CAD were more female, more frequent with a medical history of atrial fibrillation, and had a lower hemoglobin level at admission. However, they were less likely to have atherosclerotic cardiovascular disease (ASCVD) risk factors such as hypertension or smoking. 124 cases (65.6%) of atrial septal defect were the commonest comorbidity with CAD. In the matched cohort, baseline characteristics between the two groups were well balanced except for the history of hypertension (56.1% vs 67.1%, p<0.001).

**Table 1.**
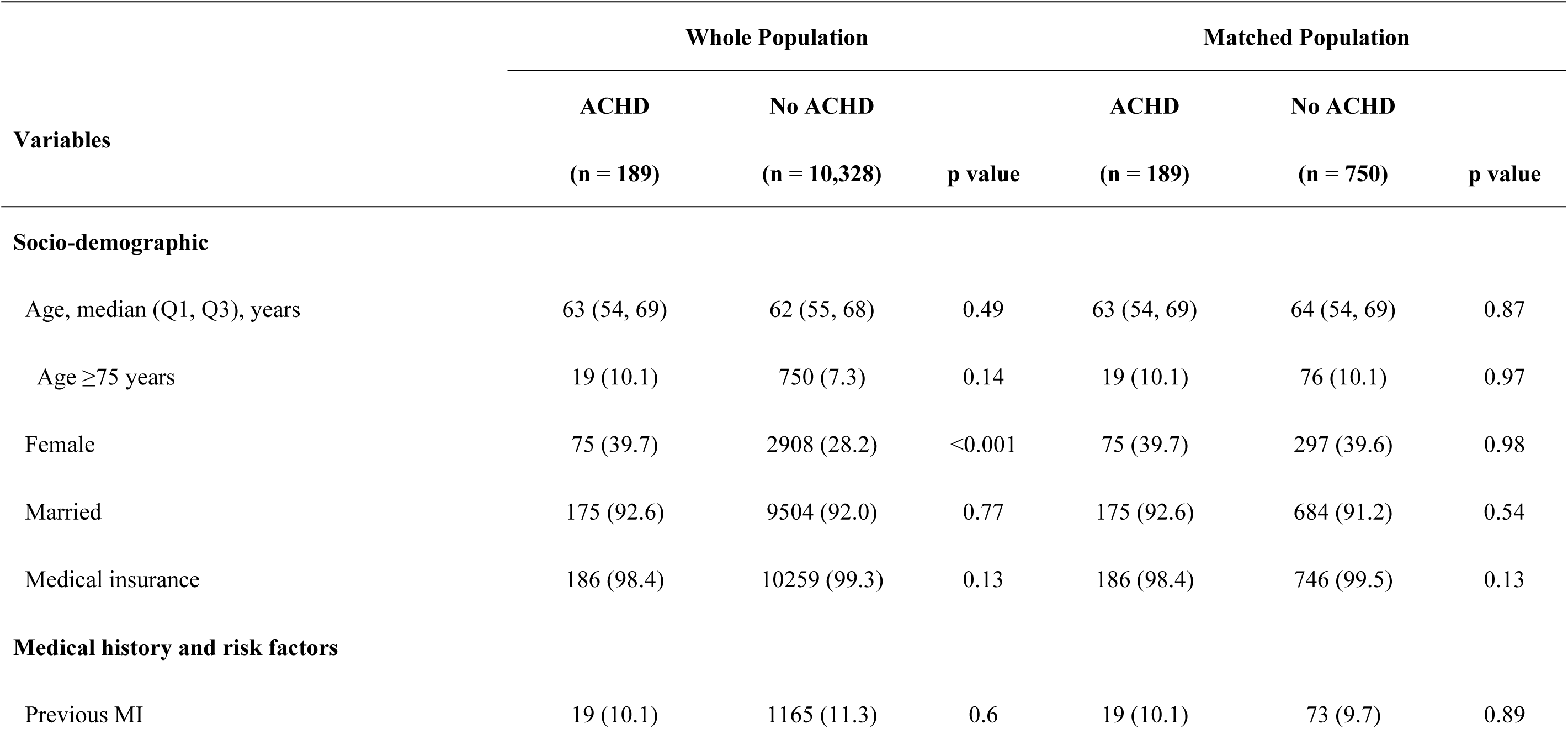
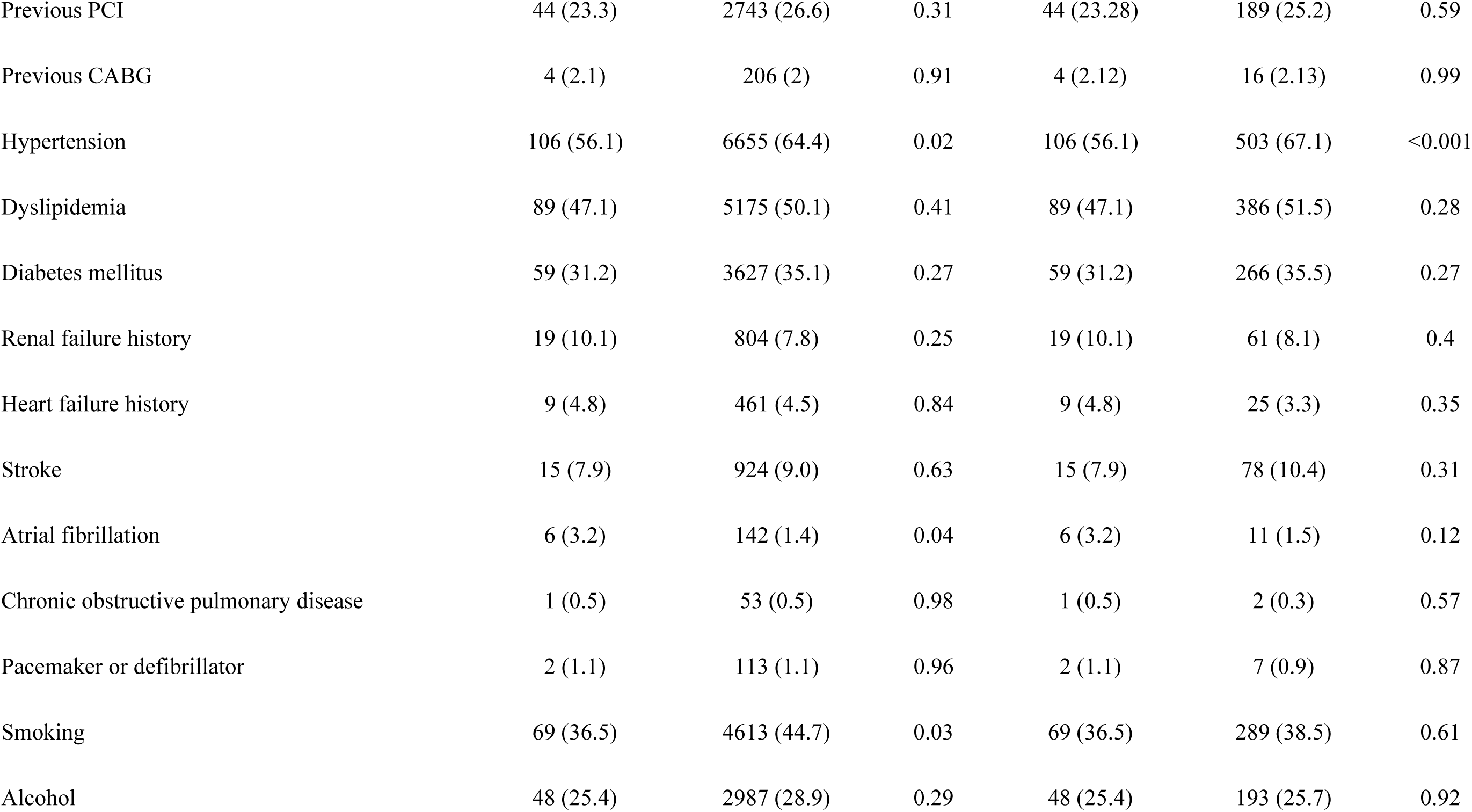
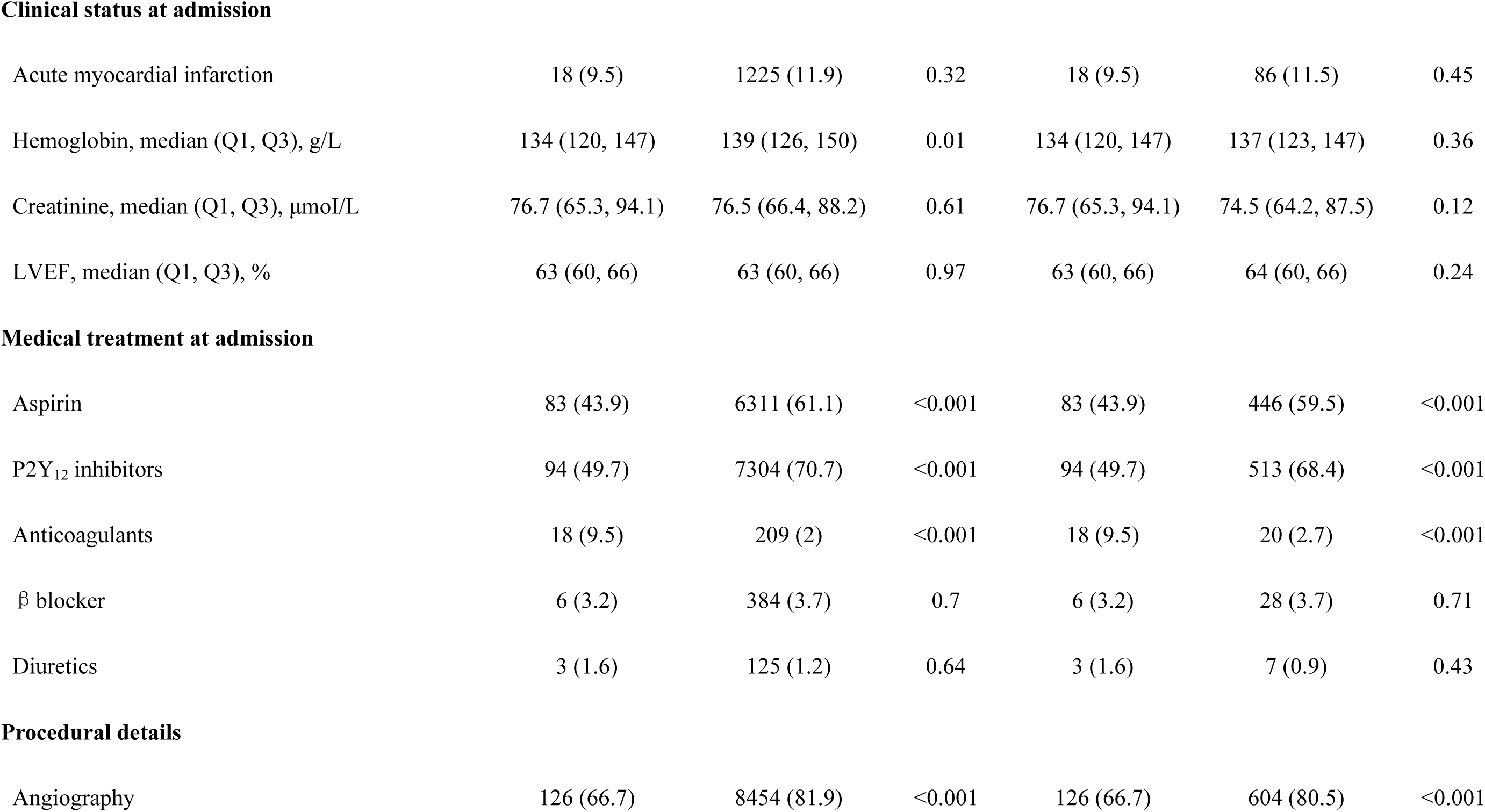
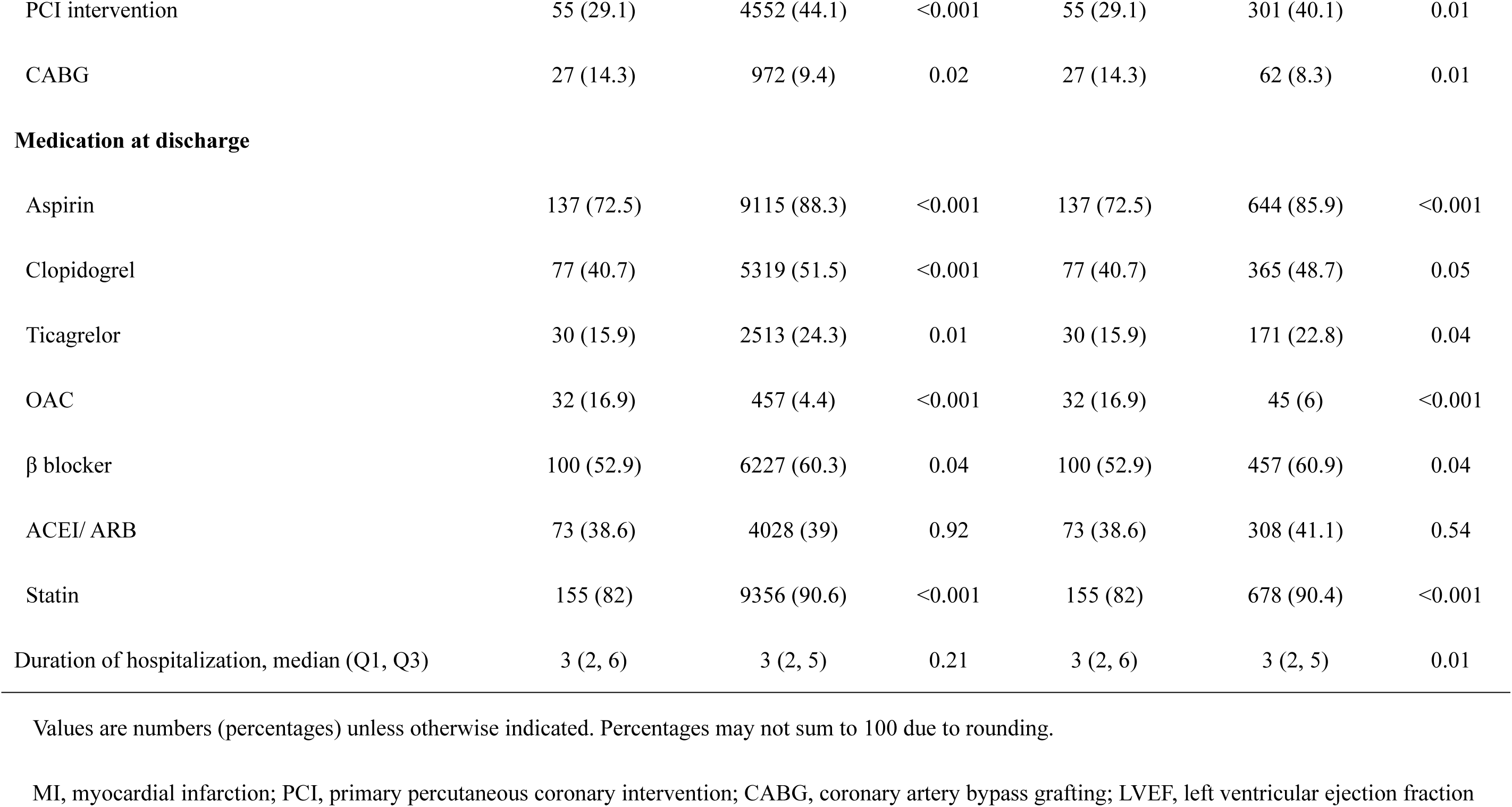
Baseline Characteristics and treatments.

| Variables | Whole Population |  |  | Matched Population |  |  |
| --- | --- | --- | --- | --- | --- | --- |
|  | ACHD | No ACHD | p value | ACHD | No ACHD | p value |
|  | (n = 189) | (n = 10,328) |  | (n = 189) | (n = 750) |  |
| Socio-demographic |  |  |  |  |  |  |
| Age, median (Q1, Q3), years | 63 (54, 69) | 62 (55, 68) | 0.49 | 63 (54, 69) | 64 (54, 69) | 0.87 |
| Age ≥75 years | 19 (10.1) | 750 (7.3) | 0.14 | 19 (10.1) | 76 (10.1) | 0.97 |
| Female | 75 (39.7) | 2908 (28.2) | <0.001 | 75 (39.7) | 297 (39.6) | 0.98 |
| Married | 175 (92.6) | 9504 (92.0) | 0.77 | 175 (92.6) | 684 (91.2) | 0.54 |
| Medical insurance | 186 (98.4) | 10259 (99.3) | 0.13 | 186 (98.4) | 746 (99.5) | 0.13 |
| Medical history and risk factors |  |  |  |  |  |  |
| Previous MI | 19 (10.1) | 1165 (11.3) | 0.6 | 19 (10.1) | 73 (9.7) | 0.89 |
| Previous PCI | 44 (23.3) | 2743 (26.6) | 0.31 | 44 (23.28) | 189 (25.2) | 0.59 |
| Previous CABG | 4 (2.1) | 206 (2) | 0.91 | 4 (2.12) | 16 (2.13) | 0.99 |
| Hypertension | 106 (56.1) | 6655 (64.4) | 0.02 | 106 (56.1) | 503 (67.1) | <0.001 |
| Dyslipidemia | 89 (47.1) | 5175 (50.1) | 0.41 | 89 (47.1) | 386 (51.5) | 0.28 |
| Diabetes mellitus | 59 (31.2) | 3627 (35.1) | 0.27 | 59 (31.2) | 266 (35.5) | 0.27 |
| Renal failure history | 19 (10.1) | 804 (7.8) | 0.25 | 19 (10.1) | 61 (8.1) | 0.4 |
| Heart failure history | 9 (4.8) | 461 (4.5) | 0.84 | 9 (4.8) | 25 (3.3) | 0.35 |
| Stroke | 15 (7.9) | 924 (9.0) | 0.63 | 15 (7.9) | 78 (10.4) | 0.31 |
| Atrial fibrillation | 6 (3.2) | 142 (1.4) | 0.04 | 6 (3.2) | 11 (1.5) | 0.12 |
| Chronic obstructive pulmonary disease | 1 (0.5) | 53 (0.5) | 0.98 | 1 (0.5) | 2 (0.3) | 0.57 |
| Pacemaker or defibrillator | 2 (1.1) | 113 (1.1) | 0.96 | 2 (1.1) | 7 (0.9) | 0.87 |
| Smoking | 69 (36.5) | 4613 (44.7) | 0.03 | 69 (36.5) | 289 (38.5) | 0.61 |
| Alcohol | 48 (25.4) | 2987 (28.9) | 0.29 | 48 (25.4) | 193 (25.7) | 0.92 |
| **Clinical status at admission** |  |  |  |  |  |  |
| Acute myocardial infarction | 18 (9.5) | 1225 (11.9) | 0.32 | 18 (9.5) | 86 (11.5) | 0.45 |
| Hemoglobin, median (Q1, Q3), g/L | 134 (120, 147) | 139 (126, 150) | 0.01 | 134 (120, 147) | 137 (123, 147) | 0.36 |
| Creatinine, median (Q1, Q3), $\mu\text{mol/L}$ | 76.7 (65.3, 94.1) | 76.5 (66.4, 88.2) | 0.61 | 76.7 (65.3, 94.1) | 74.5 (64.2, 87.5) | 0.12 |
| LVEF, median (Q1, Q3), % | 63 (60, 66) | 63 (60, 66) | 0.97 | 63 (60, 66) | 64 (60, 66) | 0.24 |
| **Medical treatment at admission** |  |  |  |  |  |  |
| Aspirin | 83 (43.9) | 6311 (61.1) | <0.001 | 83 (43.9) | 446 (59.5) | <0.001 |
| P2Y<sub>12</sub> inhibitors | 94 (49.7) | 7304 (70.7) | <0.001 | 94 (49.7) | 513 (68.4) | <0.001 |
| Anticoagulants | 18 (9.5) | 209 (2) | <0.001 | 18 (9.5) | 20 (2.7) | <0.001 |
| $\beta$ blocker | 6 (3.2) | 384 (3.7) | 0.7 | 6 (3.2) | 28 (3.7) | 0.71 |
| Diuretics | 3 (1.6) | 125 (1.2) | 0.64 | 3 (1.6) | 7 (0.9) | 0.43 |
| **Procedural details** |  |  |  |  |  |  |
| Angiography | 126 (66.7) | 8454 (81.9) | <0.001 | 126 (66.7) | 604 (80.5) | <0.001 |
| PCI intervention | 55 (29.1) | 4552 (44.1) | <0.001 | 55 (29.1) | 301 (40.1) | 0.01 |
| CABG | 27 (14.3) | 972 (9.4) | 0.02 | 27 (14.3) | 62 (8.3) | 0.01 |
| **Medication at discharge** |  |  |  |  |  |  |
| Aspirin | 137 (72.5) | 9115 (88.3) | <0.001 | 137 (72.5) | 644 (85.9) | <0.001 |
| Clopidogrel | 77 (40.7) | 5319 (51.5) | <0.001 | 77 (40.7) | 365 (48.7) | 0.05 |
| Ticagrelor | 30 (15.9) | 2513 (24.3) | 0.01 | 30 (15.9) | 171 (22.8) | 0.04 |
| OAC | 32 (16.9) | 457 (4.4) | <0.001 | 32 (16.9) | 45 (6) | <0.001 |
| β blocker | 100 (52.9) | 6227 (60.3) | 0.04 | 100 (52.9) | 457 (60.9) | 0.04 |
| ACEI/ ARB | 73 (38.6) | 4028 (39) | 0.92 | 73 (38.6) | 308 (41.1) | 0.54 |
| Statin | 155 (82) | 9356 (90.6) | <0.001 | 155 (82) | 678 (90.4) | <0.001 |
| Duration of hospitalization, median (Q1, Q3) | 3 (2, 6) | 3 (2, 5) | 0.21 | 3 (2, 6) | 3 (2, 5) | 0.01 |

Values are numbers (percentages) unless otherwise indicated. Percentages may not sum to 100 due to rounding.
MI, myocardial infarction; PCI, primary percutaneous coronary intervention; CABG, coronary artery bypass grafting; LVEF, left ventricular ejection fraction

The most frequently used thromboprophylaxis for ACHD was anticoagulant, with weak antiplatelet therapy than no ACHD group. This drug discrimination was observed both at admission and discharge. The ACHD patients underwent CABG more frequently than the no ACHD group (14.3% vs 9.4%, p=0.02), but less received PCI (29.1% vs 44.1%, p<0.001). Moreover, compared with no ACHD, the ACHD patients had a less frequent use of statin at discharge (74.6% vs 86.7%, p<0.001). Similar results were found in the matched cohort.

### In-hospital clinical outcomes

**Table 2** shows in-hospital outcomes in the two groups. In-hospital mortality before discharge was higher in patients with ACHD compared with no ACHD in the whole population (1.06% vs. 0.28%, p=0.05) and matched population (1.06% vs. 0.13%, p=0.04). The overall incidence of in-hospital cardiovascular death was 0.28% (29 cases); the incidence of cardiovascular death was higher in patients with ACHD than in those without (1.06% vs. 0.26%; p=0.04). Compared with no ACHD, ACHD group has an increased risk of in-hospital stroke (3.17% vs 0.93%, p<0.001), ischemic stroke and intracerebral hemorrhage; while the difference in ischemic stroke was no longer significant in the matched population. The incidence of lung infection during hospitalization was similar in the two groups (0.53% vs. 0.44%, p=0.85).

**Table 2.** Incidence of In-hospital Clinical Outcomes between study groups.

| Outcomes | Whole population |  |  | Matched population |  |  |
| --- | --- | --- | --- | --- | --- | --- |
|  | ACHD | No ACHD | p value | ACHD | No ACHD | p value |
|  | (n = 189) | (n = 10,328) |  | (n = 189) | (n = 750) |  |
| Death | 2 (1.06) | 29 (0.28) | 0.05 | 2 (1.06) | 1 (0.13) | 0.04 |
| Cardiovascular death | 2 (1.06) | 27 (0.26) | 0.04 | 2 (1.06) | 1 (0.13) | 0.04 |
| Stroke | 6 (3.17) | 96 (0.93) | <0.001 | 6 (3.17) | 9 (1.2) | 0.05 |
| Ischemic stroke | 5 (2.65) | 93 (0.9) | 0.01 | 5 (2.65) | 9 (1.2) | 0.14 |
| Intracerebral hemorrhage | 1 (0.53) | 3 (0.03) | <0.001 | 1 (0.53) | 0 (0) | 0.05 |
| Pneumonia | 1 (0.53) | 45 (0.44) | 0.85 | 1 (0.53) | 2 (0.27) | 0.57 |

The causes of death are presented in **Figure 2**. The most common causes of death among all adults with CAD in the death cohort were acute myocardial infarction (54.8%), cardiac shock (12.9%) and cardiac arrest (9.6%). The 2 deaths among those with ACHD were due to ischemic stroke (100%).

**Figure 2.**
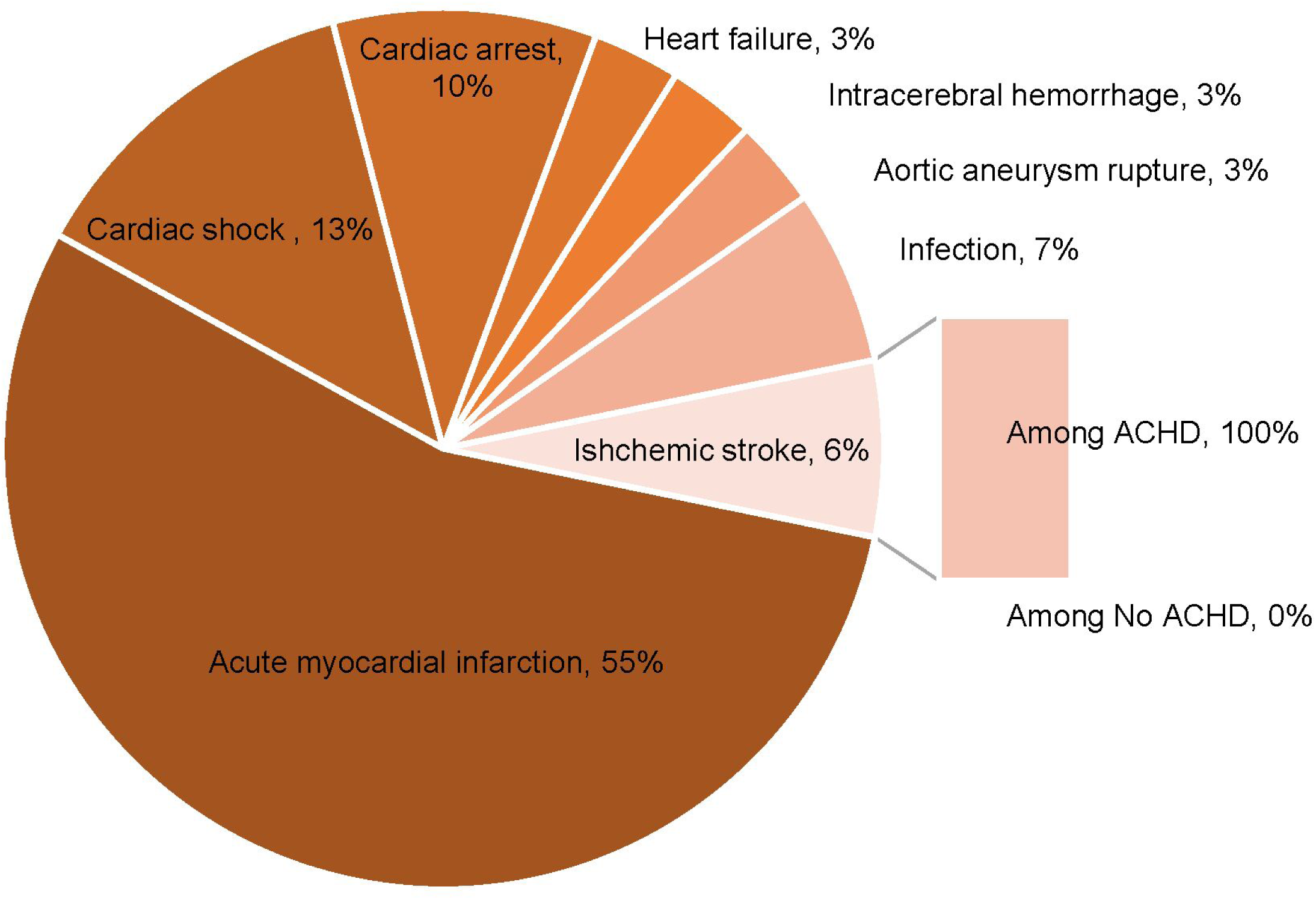
Causes of death among study cohort. ACHD, congenital heart disease

### Risk factors of death and stroke

In the logistic regression model including baseline characteristics with a statistically significant difference between groups, anemia at admission (OR: 4.57, 95% CI, 1.9–11.01), acute myocardial infarction (OR: 19.15, 95% CI, 8.77–41.82) were independent risk factors of mortality (**Table 3**). Whereas, the PCI intervention (OR: 0.38, 95% CI, 0.15–0.95) and CABG (OR: 19.15, 95% CI, 8.77–41.82) were identified as protectors of mortality. Compared with patients without ACHD, the risk of stroke markedly increased in patients with ACHD (OR: 2.89, 95% CI, 1.22–6.87). Moreover, the history of hypertension showed a strong association with in-hospital stroke (OR: 2.9, 95% CI, 1.74–4.83).

**Table 3.** Risk factors of death and strokes

|  | Death |  | Stroke |  |
| --- | --- | --- | --- | --- |
|  | OR (95% CI) | p value | OR (95% CI) | p value |
| ACHD | 3.03 (0.65–14.13) | 0.16 | 2.89 (1.22–6.87) | 0.02 |
| Female | 1.23 (0.47–3.25) | 0.67 | 1.25 (0.77–2.01) | 0.36 |
| Anemia at admission | 4.57 (1.9–11.01) | <0.001 | 1.08 (0.62–1.89) | 0.78 |
| Smoking | 1.6 (0.65–3.94) | 0.3 | 0.74 (0.45–1.2) | 0.22 |
| Hypertension | 1.98 (0.85–4.57) | 0.11 | 2.9 (1.74–4.83) | <0.001 |
| Atrial fibrillation | 2.81 (0.36–21.73) | 0.32 | N/A | N/A |
| Acute myocardial infarction | 19.15 (8.77–41.82) | <0.001 | 1.44 (0.78–2.69) | 0.24 |
| PCI intervention | 0.38 (0.15–0.95) | 0.04 | 0.32 (0.17–0.59) | <0.001 |
| CABG | 0.14 (0.04–0.5) | 0.002 | 1.09 (0.58–2.04) | 0.78 |

PCI, primary percutaneous coronary intervention; CABG, coronary artery bypass grafting

## DISCUSSION

Among 10,517 hospitalized CAD patients in the real-world cohort who were analyzed in this study, ACHD was associated with a higher rate of mortality driven by stroke complications. Furthermore, no differences were found between ACHD and no ACHD patients with regard to the risk of pneumonia.

Nowadays, the medical need in patients with CHD is shifted from survival to beyond survival^11, 12^. Truly assessing the long-term impact of CHD is mandatory for people living with chronic conditions and their caregivers^13^. Whereas consideration and overcoming above mentioned challenges of the utmost importance in guidelines, comprehensive registries with contemporary CHD data are warrant^11^. In a retrospective, a single-center database of patient older than 40 year and patent foramen ovale (PFO) in Toronto, 180 patients (84.9%) did concurrent coronary angiography with PFO disclosure^14^. A low prevalence of moderate-to-severe CAD (12.8%) was identified by coronary angiography and rare (0.6%) underwent concurrent angioplasty. A report from a standard adult cardiovascular center between April 2012 and August 2019, only 58 patients with the diagnosis of ACHD have been hospitalized^15^. Among them, only 6 (10.3%) were comorbid CAD. Lately, a national wide registry from Dutch demonstrated an increased CAD risk in ACHD patients compared to the general population, especially in youth but not in women older than 65 years or men older than 50 years^16^. However, these data cannot reach an established opinion regarding the accurate rate of comorbid CAD with ACHD, because of the diversity of the studied population. In our database using routine medical data sources, a real-world cohort of coronary artery disease, showed 1.8% CAD patients with ACHD. As the high incidence of mortality was observed in the ACHD and CAD population, which highlights the complexity mechanisms other than atherosclerosis contribute to the high mortality associated with CHD in this population. Although our observational study might be influenced by potentially unmeasured confounding variables, the data including conversational, interventional, and surgical data shed light on this important remark.

Recently, a higher rate of recurrent ischemic events was observed in ACHD population, especially of neurocognitive complications^17, 18^. Analysis from large cohort of adults with mild-to-moderate CHD of UK-biobank demonstrated measurable neurocognitive deficits. These findings indicated a transition cerebro- and cardiovascular process for CHD individuals moving from pediatric to adult^18^. In a patient-level pooled analysis of a randomized trial, young adults with cryptogenic strokes of undetermined source (ESUS) showed a low rate of recurrence stroke than older adults with ESUS^4^. Whereas PFO was detected in 50% of participants during follow-up, which may represent an incidental risk factor for stroke occurrence. Shah P, et al. systematic reviewed 386 cases with a thrombus straddling a PFO from 359 publications^3^. In their research, acute myocardial infarction but not ischemic stroke was associated with an 8-fold increased risk of in-hospital mortality. Analysis of individual patient data comprised of observational and randomized studies focused on PFO population. These results indicated atrial septal aneurysm as an important predictor of recurrent stroke, suggesting the need for risk stratification and procedural intervention in this population. In line with the previous studies, an increased rate of stroke was observed in ACHD group, suggesting the need for future studies to improve acknowledge of underlying stroke mechanisms in this special population. A priori, it may have been hypothesized that surgical impairment would reduce the rate of acute thrombotic complications which resulting ischemic events. The on-going trials (NCT03690518, NCT04902768, NCT04288596) will provide firmer information on this question^19^.

In our study, comorbid ACHD and history of hypertension were associated with a higher rate of in-hospital stroke. A report from CHD centers in North Carolina from 2008 to 2013 including 629 CHD patients dead with age at over 20 years, showed causes of death differed by lesion severity^20^. ACHD patients with severe lesions commonly died from underlying CHD, whereas those with non-severe CHD more commonly died from ischemic heart disease or malignancy. Hypertension and hyperlipidemia were prevalent at high rates among ACHD, indicating longitudinally medical care for ACHD should focus on traditional ASCVD risk factors. Latterly, Swedish National Patient Register identified 17,189 ACHD patients and 180,131 matched controls describing the risk of myocardial infarction (MI) in ACHD^21^. In line with the national report, our data shows ACHD patients died commonly due to new-onset heart failure, or death after the index MI. Recognizing and managing the modifiable ASCVD risk factors are crucial to reduce morbidity and mortality in this special population. More detailed studies of these links are warranted to establish potential causality and pathogenesis ought to investigate the predictor and outcomes of ischemic event including MI or stroke in ACHD population.

Several important limitations of this study should be noted. Consisted with all observational studies, inherent limitations and residual confounding exist, our results should be considered as hypothesis-generating. Nevertheless, the study provides pragmatic data reflecting contemporary management in ACHD patients with CAD in China. Second, we did not include variables related to invasive extracorporeal circulation in the multivariable logistic regression models exploring the risk factor of stroke, because of overfitting effect due to the small number of events. Third, we did not have detailed information on anticoagulation in the periprocedural period, which may represent a potential bias against the main objective of our study. Fourth, there were no follow-up data about the influence of stroke on mortality, disability, and quality of life. Despite the limitations, this is a pool of data picturing the view of ACHD in patients with CAD in the contemporary era.

## CONCLUSIONS

Among CAD patients with comorbid ACHD, non-severe CHD was common and ACHD was associated with an increased risk of developing stroke or death. Longitudinal study on modifiable ASCVD risk factors in this special population are awaited.

## Data Availability

Requests for the data underlying this article can be made to the corresponding author.

## CONTRIBUTION

HL and YY contributed to the manuscript draft and analysis; YY, SQ and HZ contributed to the manuscript development; SW, YX and HZ were responsible for overseeing and monitoring the implementation of the registry; all authors critically reviewed the manuscript and approved the final version.

